# WAVES (Web-based tool for Analysis and Visualization of Environmental Samples) – a web application for visualization of wastewater pathogen sequencing results

**DOI:** 10.1101/2022.05.31.22275831

**Authors:** Petr Triska, Fabian Amman, Lukas Endler, Andreas Bergthaler

## Abstract

Environmental monitoring of pathogens provides an accurate and timely source of information for public health authorities and policymakers. In the last two years, wastewater sequencing proved to be an effective way of detection and quantification of SARS-CoV-2 variants circulating in population. Wastewater sequencing produces substantial amounts of geographical and genomic data. Proper visualization of spatial and temporal patterns in this data is crucial for the assessment of the epidemiological situation and forecasting. Here, we present a web-based dashboard application for visualization and analysis of data obtained from sequencing of environmental samples. The dashboard provides multi-layered visualization of geographical and genomic data. It allows to display frequencies of detected pathogen variants as well as individual mutation frequencies. The features of WAVES for early tracking and detection of novel variants in the wastewater are demonstrated in an example of BA.1 variant and the signature Spike mutation S:E484A. WAVES dashboard is easily customized through the editable configuration file and can be used for different types of pathogens and environmental samples.

**Availability:** WAVES source code is freely available at https://github.com/ptriska/WavesDash under MIT license.

## Introduction

Wastewater monitoring has been an effective method of pathogen surveillance during the SARS-CoV-2 pandemic. In countries with developed wastewater treatment infrastructure, wastewater monitoring can provide sensitive and large-scale pathogen monitoring on the population level [1]. Genome sequencing of the SARS-CoV-2 virus in wastewater allows for reliable quantification of variants detected in the wastewater sample and even detection of novel mutations.

Pathogen sequencing from wastewater results in large amounts of spatial temporal data that can be challenging to visualize and present to policymakers and the public. Here, we describe software designed for detailed visualization of the results of wastewater genome sequencing within the synoptical dashboard WAVES (Web-based tool for Analysis and Visualization of Environmental Samples). Analysis of genomic variants in environmental samples is largely based on frequencies of variant-specific alleles and quantification of variants. Spatial and temporal patterns in variant frequencies are particularly important for public health agencies as they help to identify introduction points of novel variants and to spot local outbreaks. Our visualization tool makes finding these patterns easier. We can illustrate this with the example of initial spread of BA.1 variant in Austria in December 2021. While the BA.1 variant was not reliably detected (frequency > 10%) in wastewater in Austria until 19^th^ December 2021, the characteristic Spike mutation S:E484A was detected at a high frequency more than two weeks earlier in sample from Leoben (Figure 1, from raw data included in [1]). WAVES dashboard works as a web application. It does not require a server deployment and can be run locally on any operating system which supports Docker. A new instance of the application is initiated for every user and the application itself is stateless. The application comes with the Docker container [2] and can be hosted on a local machine, or it can be easily deployed in cloud services such as Azure or Amazon Web Services. The application is configured through the configuration file and the metadata files. By changing the configuration file and the genome map file, the application can be customized for use in diverse types of projects where genomic and spatial temporal data are analyzed together.

**Figure 1:**
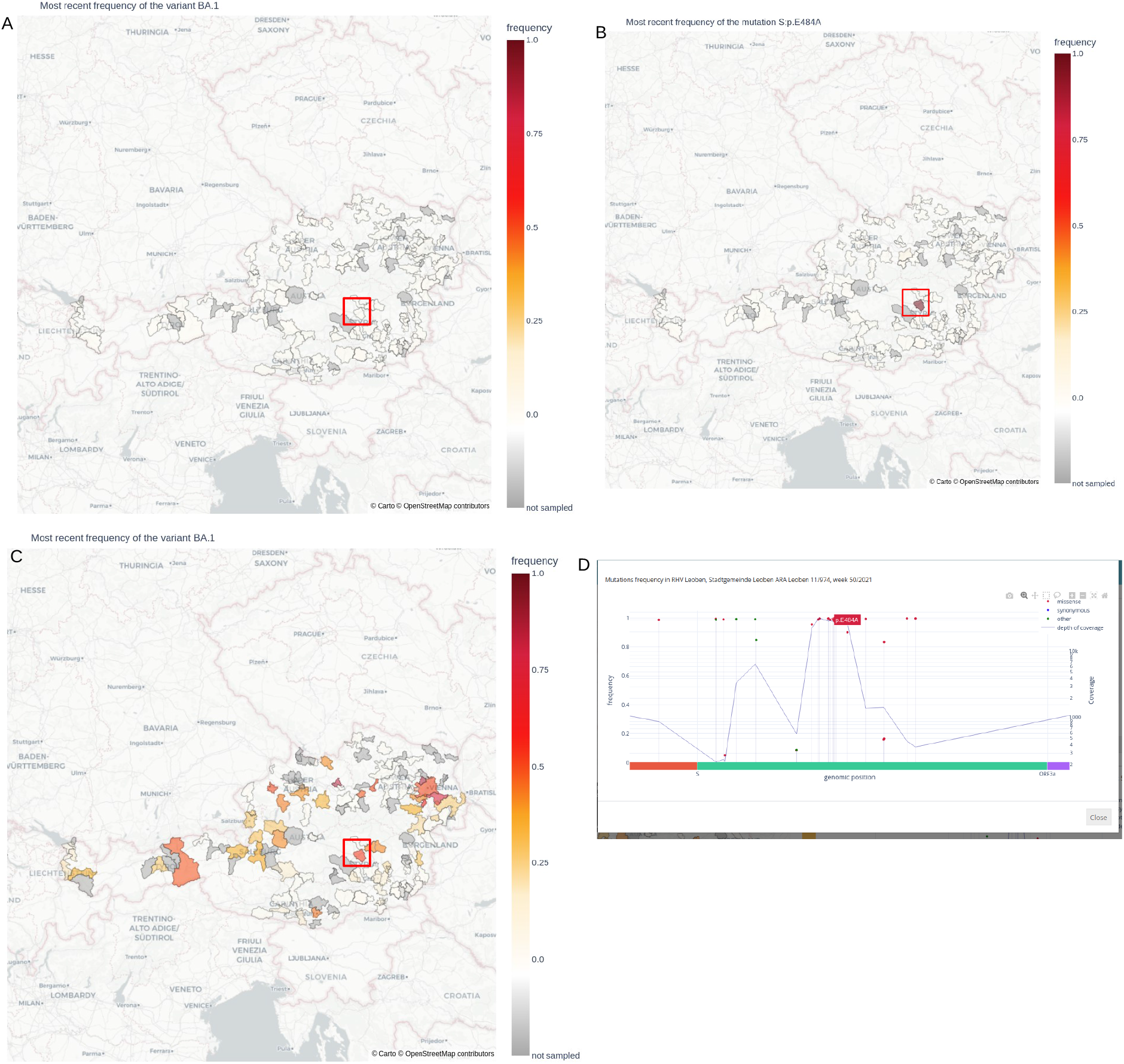
Example of use of the multi-layered data visualization with the application WAVES. Subplots A and B show the frequency of detected BA.1 variant (subplot A) and the BA.1-characteristic mutation S:E484A (subplot B) in the time period from the 1^st^ to the 15^th^ of December 2022 in Austria. While the BA.1 variant was not reliably detected in this period, the signature mutation S:E484A was already detected in the wastewater treatment plant Leoben. Two weeks later (subplot C) the Omicron variant is detected at a high level in Leoben. The detected mutation is shown in the allele frequency plot (subplot D).

The application is written in Python 3.6. [3]. The backend uses the pandas [4] and Datatable [5] packages for parsing and processing the data. The frontend is based on Plotly and Plotly Dash [6] packages for visualization of the data and for handling of user callbacks. The GitHub repository with the code can be accessed at https://github.com/ptriska/WavesDash. The application was used to produce interactive supplementary data for the recent paper by Amman et al. [1]: http://www.sarscov2-austria.org/cemm/austrian-sars-cov-2-ww-dashboard/.

### Application description

#### Backend description

WAVES integrates two types of data: frequencies of detected variants within individual samples, and the frequency of detected mutations within the sample. The application does not calculate either of those metrics and data must be supplied in the form of tab-separated files (tsv). Parsing of the tsv files is based on the Datatable package [5]. Parsed Datatable objects are then converted to pandas dataframes. Detailed description of formatting of the input files in the readme manual located in the GitHub repository.

The application parses the input files when the session is initiated. To make usage of the application smoother, the application generates pre-computed data files during the first start-up. The pre-computation performs a basic QC on the input files (removal of duplicates, indexing by sampling date and sorting of the index). In the next step, the variant frequencies are aggregated by week of the year. The pre-computed files are saved in the working folder as variant_freq_processed.tsv (variant frequencies) and allele_freq_processed.tsv (frequencies of mutations).

The update of data is done by simply removing the original data files, including the pre-computed files, and replacing them with new files. The application does not need to be restarted.

#### Frontend description

On the frontend side, WAVES consists of three vertical panels (Figure 2). On the left-hand side a divide with an interactive two-layered map is located. The first layer is a CARTO background map [7]. The center point of the map and the zoom level are specified in the configuration file. The second layer consists of outlined sampling areas, which would correspond to the respective catchment areas of a given wastewater treatment plant and is defined in the user-supplied geojson file. The sampling areas are colored in choropleth style. This can be either on qualitatively (showing the most frequent variant), or quantitatively (showing the frequency of a variant or a mutation). Outlined sampling areas are interactive. When an area is selected by clicking, a plot with sequencing results for selected area will appear in the central divide. Alternatively, when a sampling area is selected from the drop-down menu in the filtering divide, the map will zoom in to show the selected area. The map also allows a time lapse mode, in which the epidemiological situation on the map is displayed in steps corresponding to the sampling time points. The time lapse mode requires considerable amount of memory, especially when displaying long time series. For large data sets we recommend using geojson file with reduced resolution. Reduced geojson can be specified in the configuration file.

**Figure 2:**
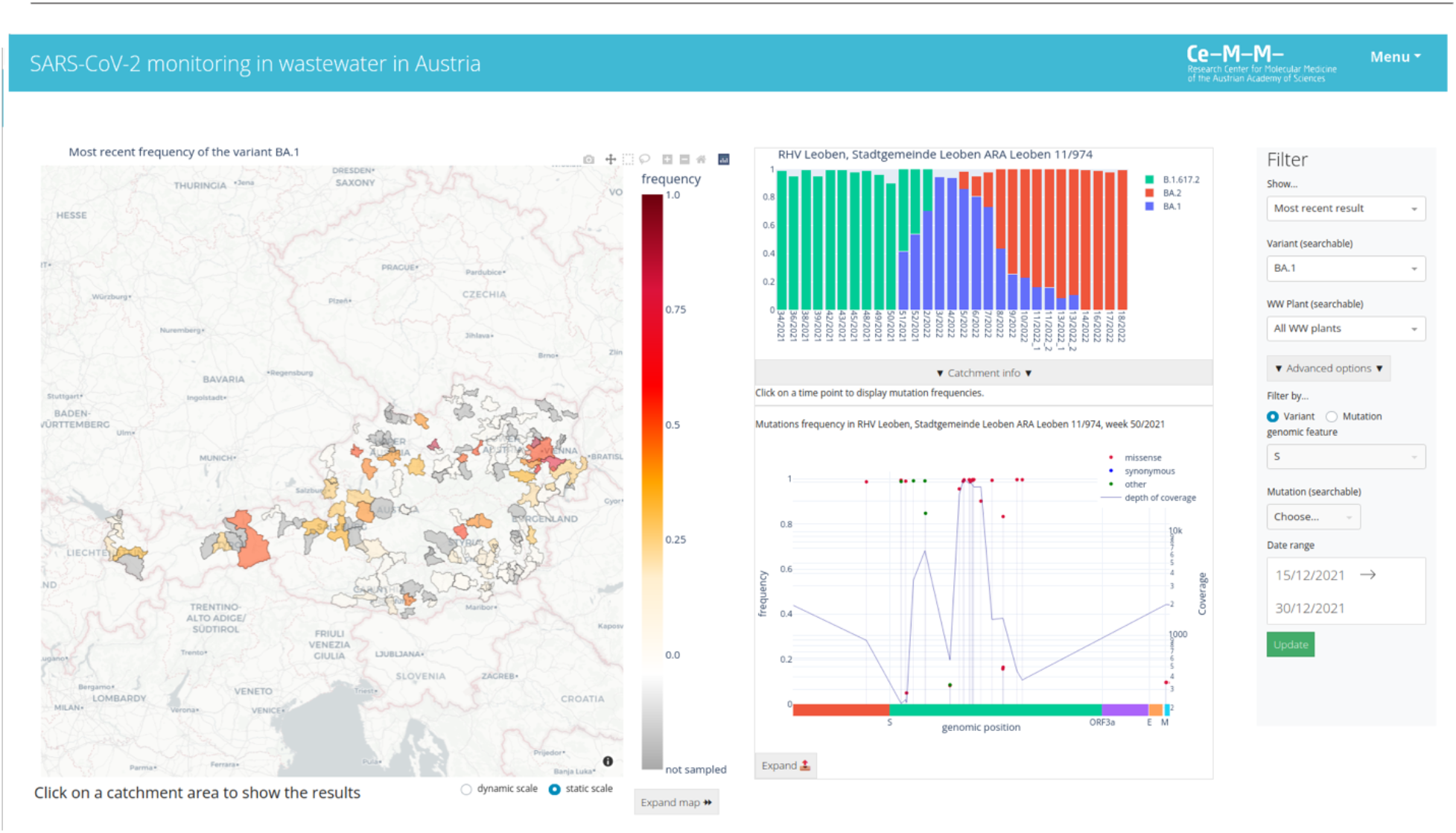
Layout of the application WAVES. A) Interactive map divide. B) Composition of detected variants at a given wastewater treatment plant. C) Allele frequencies of a given sample, displayed in a genome map.

The central divide contains two plots. The upper plot is a stacked plot showing relative frequency of each detected variant in a given sampling period. Upon selecting a sampling period by clicking at corresponding bar, a plot showing mutation frequency within a selected sample is loaded. The frequency of mutations is projected on the y-axis, while the x-axis is a genomic position of the mutation. The mutation frequency plot with the genome map can be expanded into a separate window for better readability.

The right-hand panel allows for subsetting and filtering of displayed data. Filtering is performed by selecting from drop-down menus. Filtering options contained in the drop-down forms are based on parsed data.

#### Installation

WAVES is designed to allow easy customization and deployment. Before the application is started, the user must provide a configuration file (the template of the configuration file is present in the GitHub repository), input files in the csv format, and a geojson file with geographic coordinates of sampling areas. The exact format of input files is described in the readme manual.

The application is designed to run on python 3.6+ and on Linux operating system. It uses a number of python packages that need to be installed on the system prior to the start of the application. In order to simplify the installation, we provide a pre-built Docker image containing the whole software stack. For those who prefer to build the Docker image by themselves, we provide a Docker file located in the GitHub repository.

The application starts a Flask web server which is by default accessible at localhost port 8050. The port can be changed in the configuration file.

## Conclusion

Our web-based application WAVES provides a fast and effortless way of visualization and analysis of results of the wastewater sequencing. The interaction with WAVES does not require any computational knowledge and has an intuitive user interface. It can be customized through the configuration file and can be adjusted also for other pathogens than SARS-CoV-2. The installation and deployment of WAVES is simple via the Docker container.

## Data Availability

All data produced in the present work are contained in the manuscript and available online in a Github repository.

https://github.com/ptriska/WavesDash

